# Selection of homemade mask materials for preventing transmission of COVID-19: a laboratory study

**DOI:** 10.1101/2020.05.06.20093021

**Authors:** Dijia Wang, Yanjun You, Xiaoli Zhou, Zhiyong Zong, Hao Huang, Hui Zhang, Xin Yong, Yifan Cheng, Liu Yang, Qiong Guo, Youlin Long, Yan Liu, Jin Huang, Liang Du

**Author notes:** (JH); (LD). These authors contributed equally to this work (Dijia Wang, Yanjun You).

## Abstract

The Coronavirus Disease 2019 (COVID-19) has swept the whole world with high mortality. Since droplet transmission is the main route of transmission, wearing a mask serves as a crucial preventive measure. However, the virus has spread quite quickly, causing severe mask shortage. Finding alternative materials for homemade masks while ensuring the significant performance indicators will help alleviate the shortage of masks. Referring to the national standard for the “Surgical Mask” of China, 17 materials to be selected for homemade masks were tested in four key indicators: pressure difference, particle filtration efficiency, bacterial filtration efficiency and resistance to surface wetting. Eleven single-layer materials met the standard of pressure difference (≤49 Pa), of which 3 met the standard of resistance to surface wetting (≥3), 1 met the standard of particle filtration efficiency (≥30%), but none met the standard of bacterial filtration efficiency (≥95%). Based on the testing results of single-layer materials, fifteen combinations of paired materials were tested. The results showed that three double-layer materials including double-layer medical non-woven fabric, medical non-woven fabric plus non-woven shopping bag, and medical non-woven fabric plus granular tea towel could meet all the standards of pressure difference, particle filtration efficiency, and resistance to surface wetting, and were close to the standard of the bacterial filtration efficiency. In conclusion, if resources are severely lacking and medical masks cannot be obtained, homemade masks using available materials, based on the results of this study, can minimize the chance of infection to the maximum extent.

## Introduction

In December 2019, the Coronavirus Disease 2019 (COVID-19) outbreak occurred in the city of Wuhan, Hubei province. Up to April 12, 2020, the outbreak has hit all provinces in China and 210 countries across the globe(1), which was declared as a Public Health Emergency of International Concern (PHEIC) by the World Health Organization (WHO) (2). Droplet transmission is the main routes of COVID-19 transmission. Most guidelines(3-5) recommend the use of masks to prevent droplet transmission, hence wearing a mask is one of the most important preventive measure. MacIntyre et al.(6) showed that adherence to masks significantly reduces the risk of influenza infection (HR=0.26,95%CI 0.09-0.77). Brienen et al.(7) showed that population-wide use of face masks could make an important contribution in delaying an influenza pandemic. But during the prediction period in China (from 20 Jan 2020 to 30 Jun 2020), the largest daily facemask shortages were predicted to be 589.5, 49.3, and 37.5 million in each of the three scenarios, respectively(8). Under the current global pandemic situation, the shortage of masks is still severe. Fisher et al(9), Viscusi et al.(10) have explored methods to alleviate the shortage of masks through reuse after disinfection and prolonged use time. However, with the increase of repeated uses and prolonged use time, the protective effectiveness has significantly reduced(9). Van der Sande et al.(11) have indicated that the protective factor of surgical masks was 4.1-5.3, while the protective factor of homemade masks was 2.2-2.5, which could reduce the respiratory infections of the population to a certain extent. Davies et al.(12) reported eight kinds of materials such as T-shirts, vacuum cleaner bag, tea cloth and pillowcases, significantly reduced the number of microorganisms expelled, although the surgical mask was three times more effective in blocking transmission than the homemade mask. Therefore, homemade masks using civilian materials is of great value in extreme cases of masks shortage.

The US Centers for Disease Control and Prevention advised residents to make cloth masks on their own to slow the spread of the virus on April 9(13), and the National Health Commission for Disease Control issued the “Notice on Printing and Distributing Technical Guidelines for the Selection and Use of Masks for the Prevention of COVID-19 Infections in Different Populations” on February 5, 2020, in which advises home residents, diaspora residents, outdoor activists, and low-risk groups to voluntarily wear masks(14), but lacking guidance on how to select materials scientifically. Inappropriate selection of masks may increase the chance of infection due to failing to play a protective role. Therefore, the study aims to combine the comprehensive literature and expert advice to screen the materials of homemade masks with good accessibility, then through laboratory performance testing, materials suitable for homemade masks are selected to cope with the shortage of medical masks and to protect against respiratory infectious diseases, so as to provide some references for decision-makers.

## Materials and methods

### Selection of homemade mask materials

We searched the PubMed and EMBASE databases systematically and obtained 6 studies (11, 12, 15-18)on civilian homemade mask materials under the epidemic of H5N1 and SARS, including T-shirts, scarves, tea towels, pillowcases, antibacterial pillowcases, vacuum cleaner dust bags, linen, silk, etc(S1 Table). Then, an expert consensus meeting involved eight experts in related fields including materials (2 people), nursing decontamination (2 people), evidence-based medicine and clinical epidemiology (2 people), and hospital infection management (2 people) was held to determined candidate materials for laboratoral testing. Finally, seventeen candidate materials were selected for laboratory testing, including T-shirt, fleece sweater, outdoor jacket, down jacket, sun-protective clothing, jeans, hairy tea towel, granular tea towel, non-woven fabrics shopping bag, vacuum cleaner dust bag, diaper, sanitary pad, non-woven shopping bag, vacuum cleaner bag, pillowcase A (40s × 40s air-jet down-proof fabric), pillowcase B (60s × 60s jet satin), pillowcase C (80s × 60s jet satin), medical non-woven fabric, and medical medical gauze(S2 File). Furuhashi(19) showed that the disposable mask made of glass fiber mat combined with non-woven fabric proved to be high in performance with a B.F.E. of 98.1%-99.4%. As medical device packaging materials, medical non-woven fabrics are widely applied in the field of medical device packaging owing to their high antibacterial properties and strong air permeability(20). Because it is similar to the material of medical masks, Chinese medical staff used it as a homemade mask material to improve the shortage of masks. We combined with the expertise of experts in the relevant fields and years of work experience, decided to include it in the study. Medical non-woven fabrics and medical medical gauze were obtained from the Sterilization and Supply Center of West China Hospital, and the remaining materials were purchased through malls and supermarkets(Table 1).

**Table 1.** Selected candidate materials for homemade masks.

| Material | Source | Brand | Fiber composition |
| --- | --- | --- | --- |
| T-shirt | Mall | Uniqlo | 100% cotton |
| Fleece sweater | Mall | Uniqlo | 100% cotton |
| Outdoor jacket | Mall | Decathlon | 100% polyurethane |
| Down jacket | Mall | Decathlon | 100% polyurethane |
| Sun-protective clothing | Mall | Decathlon | 100% polyester |
| Jeans | Mall | Uniqlo | 98% cotton/2% polyurethane |
| Hairy tea towel | Supermarket | Maryya | 80% polyester/20% nylon |
| Granular tea towel | Supermarket | Maryya | 80% polyester/20% nylon |
| Non-woven shopping bag | Mall | Eusu | 100% polypropylene |
| Vacuum cleaner bag | Electronic business<br>platform(Jingdong) | Dmy | 100% polyethylene-vinyl acetate |
| Diaper | Supermarket | Elderjoy | Non-woven etc |
| Sanitary pad | Supermarket | Whisper | Non-woven etc |
| Pillowcase A | Hospital | Nantong<br>Aokai | 40s×40s Air-jet down-proof fabric |
| Pillowcase B | Hospital | Nantong<br>Aokai | 60s × 60s Jet satin |
| Pillowcase C | Hospital | Nantong<br>Aokai | 80s×60s Jet satin |
| Medical non-woven fabric | Hospital | An<br>Ruiheng | 2 Layers of spunbond + 3 layers of<br>meltblown cloth |
| Medical gauze | Hospital | Rong Wei | Absorbent cotton |

### Detection Indicator

According to the national standard YY0469-2011 “Surgical Mask” and GB/T4745-2012”Textiles-Testing and evaluation for water resistance-Spray test method”(21, 22), four key indicators to detect the performance of mask materials were performed, including pressure difference, particle filtration efficiency, bacterial filtration efficiency, and resistance to surface wetting. The definitions and standards of the detection indicators are shown in Table 2(21, 22).

**Table 2.**
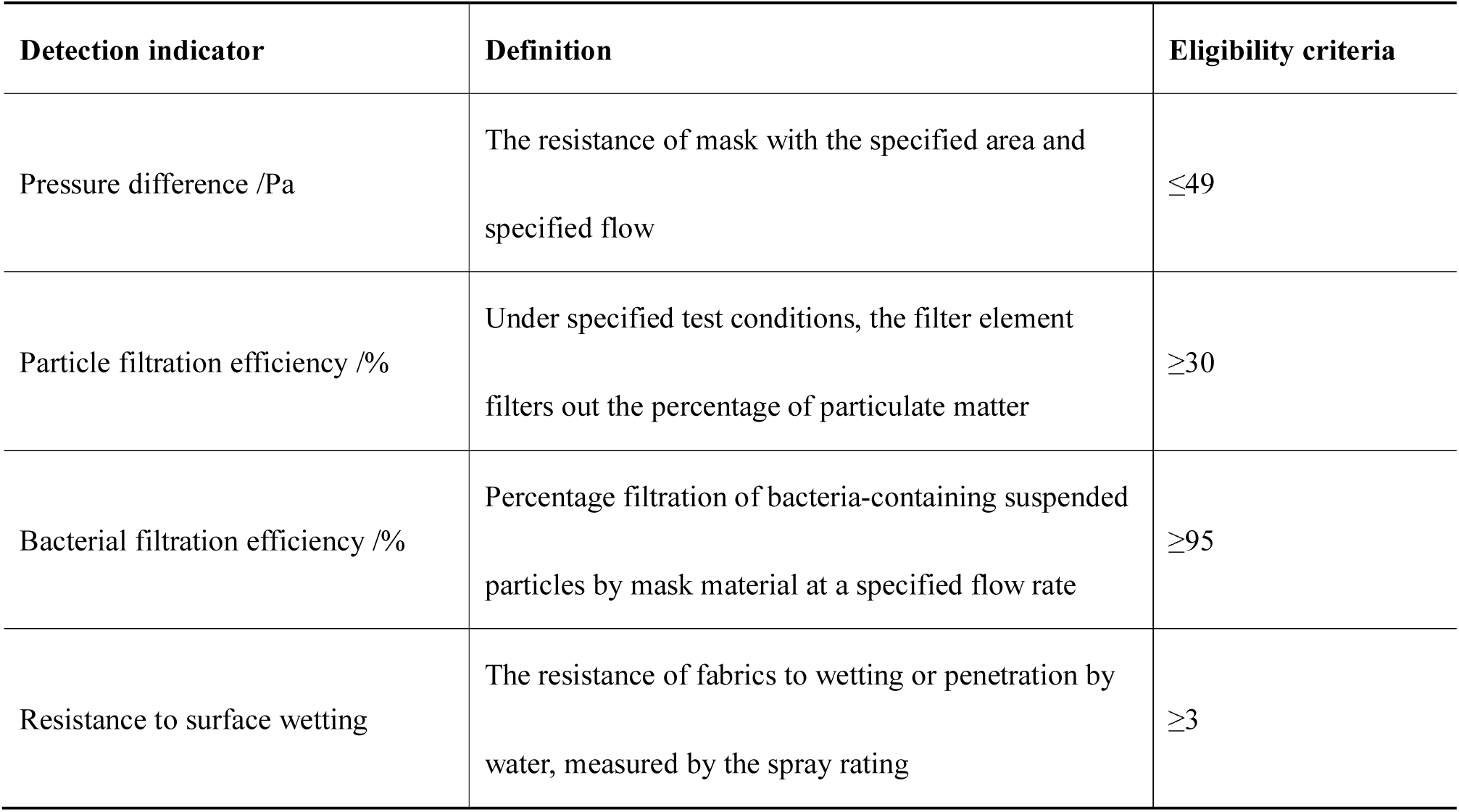
Definitions and standards of detection indicators.

| Detection indicator | Definition | Eligibility criteria |
| --- | --- | --- |
| Pressure difference /Pa | The resistance of mask with the specified area and specified flow | $\leq 49$ |
| Particle filtration efficiency /% | Under specified test conditions, the filter element filters out the percentage of particulate matter | $\geq 30$ |
| Bacterial filtration efficiency /% | Percentage filtration of bacteria-containing suspended particles by mask material at a specified flow rate | $\geq 95$ |
| Resistance to surface wetting | The resistance of fabrics to wetting or penetration by water, measured by the spray rating | $\geq 3$ |

### Experimental Methods

All materials were cut to 18*18 cm, and five samples from each material were tested by Sichuan Testing Center of Medical Devices in China. The pressure difference, particle filtration efficiency and bacterial filtration efficiency were determine by the test method stipulated in the standard of YY0469-2011 “Surgical Masks”, and resistance to surface wetting was tested in line with the test method stipulated in the standard of GB/T4745-2012 “Textiles-Testing and evaluation for water resistance-Spray test method”. The Qingdao SRP ZR-1200 medical detection instrument on surgical masks was used for pressure difference detection, with the gas flow rate at 8 L/min, the diameter of the sample test zone 25 mm, and the test area 4.9 cm^2^.

The American TSI 8130 automatic filter tester was employed for testing particle filtration efficiency. The material was first placed in an environment with a relative humidity of 85% and at 38 °C for 24 hours for pretreatment and was then sealed in an airtight container. The test was completed within 2 hours after the sample pretreatment. The test process entailed placing the pretreated material in a NaCl aerosol with a relative humidity of 30% and at 25 °C (median diameter of particle count 0.075 ± 0.020 μm), with a geometric standard deviation of the particle distribution less than 1.86 and concentration no more than 200 mg/m^3^. The gas flow rate was set to 30 L/min, and the cross-sectional area through which the air flows was 100 cm^2^.

The bacterial filtration efficiency was tested in agreement with the standard of YY0469-2011 “Surgical Mask”. The suspension of Staphylococcus aureus was prepared, followed by sterile plates placed in the A and B chambers of the Qingdao SRP ZR-1000 experimental system, with six layers in each chamber. Chamber A cavity was a positive control, and the pre-treated sample was installed in the cavity B, with the gas flow rate at 28.3 L/min, the bacterial suspension delivery time of the nebulizer 1 minute, and the operation time of sampler 2 minutes. After the test, the tryptic soy peptone agar (TSA) medium plate was incubated at 35°C for 48 hours and removed subsequently. The colony-forming units (positive pores) formed by bacterial particle aerosols were counted afterward, and the number of possible impact particles was converted in accordance with the conversion table(21), and 5 samples were tested using the same method.

The principle of resistance to surface wetting detection was to install the sample on the snap ring and place it at a 45-degree angle to the horizontal, with the center of the sample 150 mm below the nozzle, which was sprayed with 250 mL of distilled water. The spray rating was determined by comparing the appearance of the sample with the evaluation standards and pictures, using Wenzhou Darong Y(B) 813 fabric water-wetting tester as the test instrument.

The pressure difference of single-layer materials was firstly tested to exclude materials with a pressure difference over 49 Pa, and the qualified materials of the pressure difference were further tested for particle filtration efficiency, bacterial filtration efficiency and resistance to surface wetting. Because all the single-layer materials had at least one indicator that failed to meet the eligibility criteria, we, based on test results of the single-layer materials, further screened materials with qualified resistance to surface wetting as the outer layer, with the same material or other materials as the inner layer to form double-layer material, to test whether it can meet the standard. Due to the limitation of testing equipment, materials with over two layers cannot be detected thanks to their ultra-thickness, consequently, a combination of multi-layer materials was not further designed (Fig 1).

**Figure 1.**
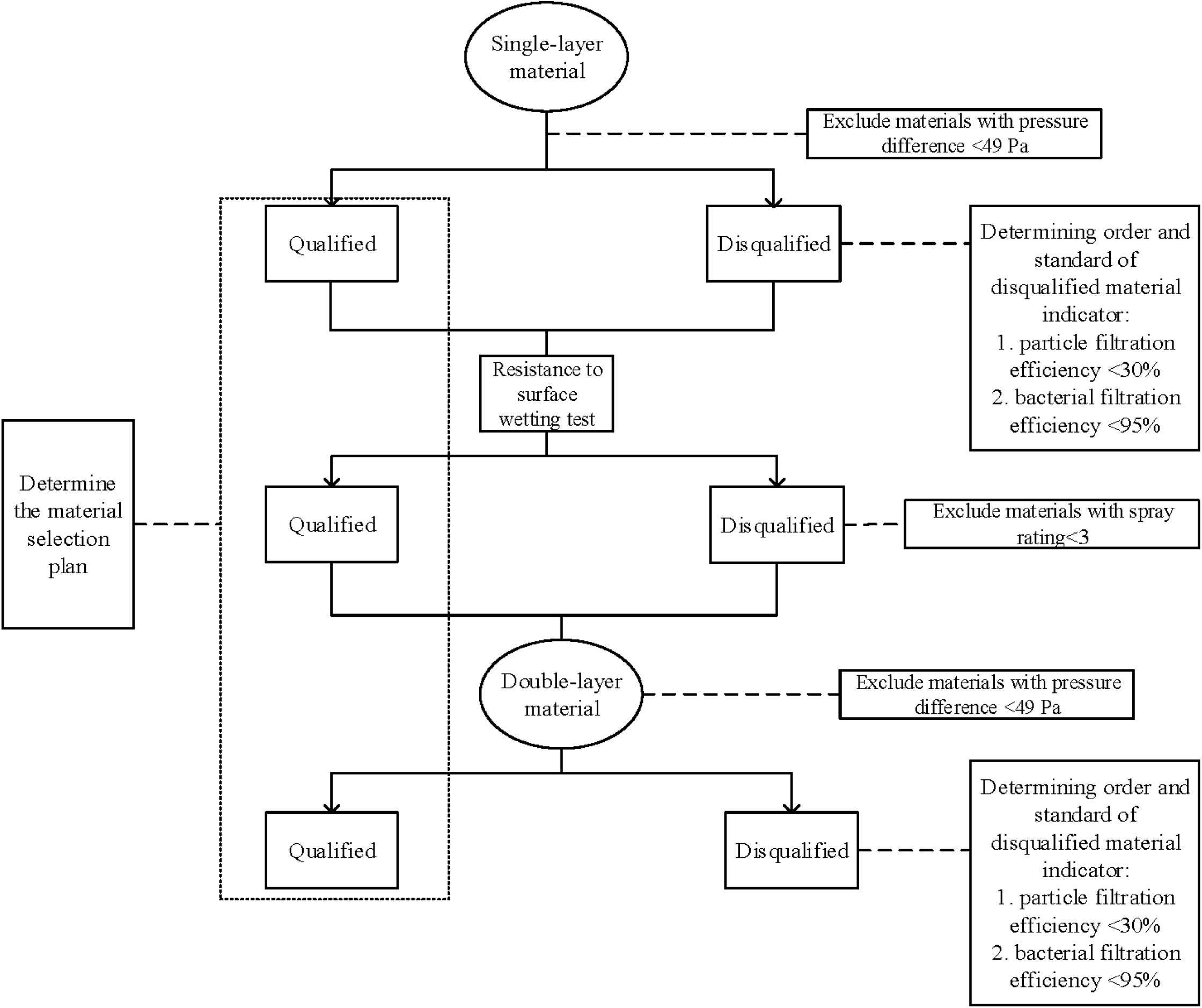

### Statistical methods

Descriptive statistical analysis of the data was conducted with the use of SPSS18.0 software. The mean, standard deviation, and 95% confidence interval (CI) of 5 samples of each material/material combination in each indicator were reported in our study.

## Results

### Test results of homemade mask materials for single-layer

The laboratory testing results showed that 11 materials for single-layer homemade masks with a pressure difference less than 49 Pa, and the order of pressure difference from small to large was as follows: granular tea towel, fleece sweater, medical gauze (4 layers), medical gauze (8 layers), non-woven shopping bag, medical gauze (12 layers), hairy tea towel, T-shirt, medical gauze (16 layers), pillowcase C, and medical non-woven fabrics. The testing results showed that a total of 7 materials for the single-layer homemade mask with a spray rating more than 3, and the order of spray rating from high to low was as follows: outdoor jacket/medical non-woven fabric, down jacket/sun protective clothing, non-woven shopping bag, vacuum cleaner bag, and fleece sweater. Only three materials met both standards of pressure difference (≤49Pa) and spray rating ≥3), including non-woven shopping bags, medical non-woven fabric, and fleece sweater (Table 3).

**Table 3.** Test results of pressure difference and spray rating for single-layer materials.

| Material | Pressure Difference |  |  | Spray Rating |  |
| --- | --- | --- | --- | --- | --- |
| | $\bar{X} \pm SD$ | 95% CI | Qualified or Not<br>( $\leq 49\text{ Pa}$ ) | Qualified or Not<br>( $\geq 3$ ) | |
| T-shirt | 15.80±1.01 | 14.54–17.06 | Yes | 0.0±0.0 | No |
| Fleece sweater | 5.86±0.42 | 5.34–6.38 | Yes | 3.4±0.5 | Yes |
| Outdoor jacket* | – | – | No | 4.4±0.5 | Yes |
| Down jacket | 125.36±0.77 | 124.40–126.32 | No | 4.2±0.4 | Yes |
| Sun-protective clothing | 125.26±1.12 | 123.87–126.65 | No | 4.2±0.4 | Yes |
| Jeans | 124.62±0.99 | 123.39–125.85 | No | 1.4±0.5 | No |
| Hairy tea towel | 13.72±0.53 | 13.06–14.38 | Yes | 1.2±0.4 | No |
| Granular tea towel | 5.72±0.13 | 5.56–5.88 | Yes | 1.4±0.5 | No |
| Non-woven shopping bag | 7.06±0.27 | 6.72–7.40 | Yes | 3.8±0.8 | Yes |
| Vacuum cleaner bag* | – | – | No | 3.8±0.4 | Yes |
| Diaper | 125.44±0.87 | 124.36–126.52 | No | 1.0±0.0 | No |
| Sanitary pad | 125.44±1.09 | 124.08–126.80 | No | 1.2±0.4 | No |
| Pillowcase A | 125.34±0.63 | 124.55–126.13 | No | 0.0±0.0 | No |
| Pillowcase B | 67.80±0.88 | 66.71–68.89 | No | 1.8±0.4 | No |
| Pillowcase C | 26.86±0.58 | 26.14–27.58 | Yes | 1.2±0.4 | No |
| Medical non-woven fabric | 35.98±1.85 | 33.68–38.28 | Yes | 4.4±0.5 | Yes |
| Medical gauze (4 layers)<sup>#</sup> | 6.02±0.64 | 5.23–6.81 | Yes | – | No |
| Medical gauze (8layers)<sup>#</sup> | 6.36±0.42 | 5.84–6.88 | Yes | – | No |
| Medical gauze (12layers)<sup>#</sup> | 8.94±0.50 | 8.32–9.56 | Yes | – | No |
| Medical gauze (16layers)<sup>#</sup> | 17.52±1.33 | 15.87–19.17 | Yes | – | No |

\* The pressure difference of outdoor jacket and vacuum cleaner bag cannot be tested caused by too high ventilation resistance.
<sup>#</sup> The resistance to surface wetting of medical gauze is not considered, given its main function is to wrap wounds and clean up bloodstains.

**Table 4.** Test results of particle filtration efficiency of materials for single-layer homemade masks with qualified pressure difference.

| Material | $\bar{X} \pm SD$ | 95%CI | Qualified or Not pass( ≥30% ) |
| --- | --- | --- | --- |
| T-shirt | 12%±1% | 11%–14% | No |
| Fleece sweater | 6%±0% | 5%–6% | No |
| Hairy tea towel | 23%±1% | 22%–24% | No |
| Granular tea towel | 12%±1% | 11%–13% | No |
| Non-woven shopping bag | 14%±2% | 12%–17% | No |
| Pillowcase C | 0%±0% | 0%–0% | No |
| Medical non-woven fabric | 42%±2% | 40%–44% | Yes |
| Medical gauze (4 layers) | 2%±0% | 2%–3% | No |
| Medical gauze (8layers) | 3%±0% | 3%–4% | No |
| Medical gauze (12layers) | 5%±0% | 5%–6% | No |
| Medical gauze (16layers) | 14%±1% | 12%–15% | No |

The particle filtration efficiency test was performed on the materials with qualified pressure difference, and it was found that only the medical non-woven fabric out of the 11 materials had a particle filtration efficiency of over 30% (mean=42%, SD=2%, 95% CI 40%-44%).

Further bacterial filtration efficiency testing results of medical non-woven fabric showed that it failed to meet the standard of more than 95% (mean=62%, SD=1%, 95% CI 60%-64%) (Table 5).

**Table 5.** Test results of bacterial filtration efficiency of materials for single-layer homemade masks with qualified pressure difference and particle filtration efficiency.

| Material | $\bar{X} \pm SD$ | 95%CI | Qualified or Not ( ≥95% ) |
| --- | --- | --- | --- |
| Medical non-woven fabric | 62%±1% | 60%–64% | No |

### Test results of homemade mask materials for double-layer

Fifteen double-layer materials were tested. The results demonstrated that there were 12 (80%) double-layer materials with a pressure difference of ≤49 Pa, with double-layer fleece sweater as the minimum pressure difference (mean=12.40, SD=1.53, 95%CI 10.50-14.30), and medical non-woven fabric plus granular tea towel as the maximum pressure difference (mean=43.52, SD=1.48, 95%CI 41.68-45.36)(Table 6).

**Table 6.**
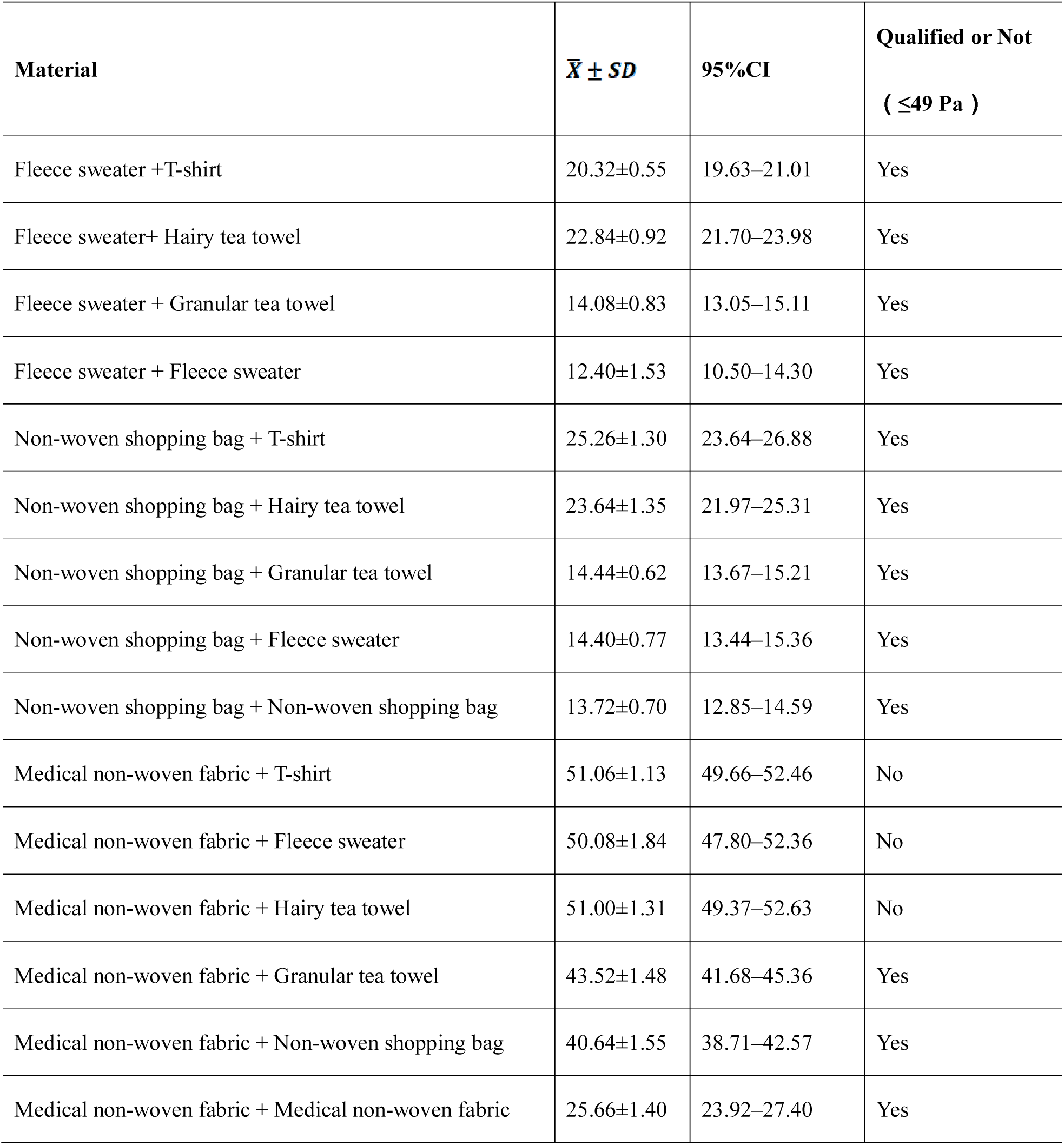
Test results of pressure difference of materials for double-layer homemade masks.

| **Material** | **$\bar{X} \pm SD$** | **95%CI** | **Qualified or Not**<br>**( $\leq 49$ Pa )** |
| --- | --- | --- | --- |
| Fleece sweater +T-shirt | 20.32±0.55 | 19.63–21.01 | Yes |
| Fleece sweater+ Hairy tea towel | 22.84±0.92 | 21.70–23.98 | Yes |
| Fleece sweater + Granular tea towel | 14.08±0.83 | 13.05–15.11 | Yes |
| Fleece sweater + Fleece sweater | 12.40±1.53 | 10.50–14.30 | Yes |
| Non-woven shopping bag + T-shirt | 25.26±1.30 | 23.64–26.88 | Yes |
| Non-woven shopping bag + Hairy tea towel | 23.64±1.35 | 21.97–25.31 | Yes |
| Non-woven shopping bag + Granular tea towel | 14.44±0.62 | 13.67–15.21 | Yes |
| Non-woven shopping bag + Fleece sweater | 14.40±0.77 | 13.44–15.36 | Yes |
| Non-woven shopping bag + Non-woven shopping bag | 13.72±0.70 | 12.85–14.59 | Yes |
| Medical non-woven fabric + T-shirt | 51.06±1.13 | 49.66–52.46 | No |
| Medical non-woven fabric + Fleece sweater | 50.08±1.84 | 47.80–52.36 | No |
| Medical non-woven fabric + Hairy tea towel | 51.00±1.31 | 49.37–52.63 | No |
| Medical non-woven fabric + Granular tea towel | 43.52±1.48 | 41.68–45.36 | Yes |
| Medical non-woven fabric + Non-woven shopping bag | 40.64±1.55 | 38.71–42.57 | Yes |
| Medical non-woven fabric + Medical non-woven fabric | 25.66±1.40 | 23.92–27.40 | Yes |

Of the 12 double-layer materials which met the standard of pressure difference, seven (58.3%) double-layer material had a particle filtration efficiency of more than 30%. The particle filtration efficiency of the fleece sweater plus hairy tea towel was more than 50%, nearly equal to that of double-layer medical non-woven fabric (Table 7).

**Table 7.** Test results of particle filtration efficiency of materials for double-layer homemade masks with qualified pressure difference.

| Material | $\bar{X} \pm SD$ | 95%CI | Qualified or Not<br>( $\geq 30\%$ ) |
| --- | --- | --- | --- |
| Fleece sweater + T-shirt | 12%±1% | 11%-13% | No |
| Fleece sweater + Hairy tea towel | 56%±1% | 54%-57% | Yes |
| Fleece sweater + Granular tea towel | 11%±1% | 10%-12% | No |
| Fleece sweater + Fleece sweater | 11%±0% | 10%-11% | No |
| Non-woven shopping bag + T-shirt | 30%±1% | 29%-31% | No |
| Non-woven shopping bag + Hairy tea towel | 47%±1% | 46%-48% | Yes |
| Non-woven shopping bag + Granular tea towel | 47%±1% | 46%-48% | Yes |
| Non-woven shopping bag + Fleece sweater | 35%±2% | 33%-37% | Yes |
| Non-woven shopping bag + Non-woven shopping bag | 19%±1% | 17%-21% | No |
| Medical non-woven fabric + Granular tea towel | 48%±1% | 48%-49% | Yes |
| Medical non-woven fabric + Non-woven shopping bag | 40%±1% | 40%-41% | Yes |
| Medical non-woven fabric + Medical non-woven fabric | 54%±1% | 53%-55% | Yes |

Concerning the bacterial filtration efficiency, none of the double-layer materials met the standard (>95%), but three double-layer materials were close to the standard, including double-layer medical non-woven fabric (mean=93%, SD=1, 95% CI 91%-95%), medical non-woven fabric plus non-woven shopping bag (mean=89%, SD=2%, 95% CI 86%-92%), and medical non-woven fabric plus granular tea towel (mean=88%, SD=4%, 95% CI 84%-92%)(Table 8).

**Table 8.** Test results of bacterial filtration efficiency of materials for double-layer homemade masks with qualified pressure difference and particle filtration efficiency.

| **Material** | **$\bar{X} \pm SD$** | **95%CI** | **Qualified or Not**<br>**( $\geq 95\%$ )** |
| --- | --- | --- | --- |
| Fleece sweater+ Hairy tea towel | 24%±3% | 21%-27% | No |
| Non-woven shopping bag + Hairy tea towel | 23%±2% | 20%-25% | No |
| Non-woven shopping bag + Granular tea towel | 17%±3% | 12%-21% | No |
| Non-woven shopping bag + Fleece sweater | 16%±1% | 15%-18% | No |
| Medical non-woven fabric + Granular tea towel | 88%±4% | 84%-92% | No |
| Medical non-woven fabric + Non-woven shopping bag | 89%±2% | 86%-92% | No |
| Medical non-woven fabric + Medical non-woven fabric | 93%±1% | 91%-95% | No |

## Discussion

Our study found that the bacterial filtration efficiency of homemade masks failed to meet the standards of surgical masks, but pressure difference and particle filtration efficiency of most materials/material combinations met the standards. For example, the pressure difference and particle filtration efficiency for medical non-woven fabric, both double-layer and single-layer, reached the standard, and the bacterial filtration efficiency of the double-layer medical non-woven fabric was close to the standard of surgical masks.

The medical non-woven fabric in our study is an SMMMS non-woven fabric composed of 2 layers of spunbond and 3 layers of meltblown fabrics. The structure of surgical masks is usually in three-layer: the outer layer is a spunbond nonwoven fabric with a water-blocking effect to prevent droplets from entering the mask; the middle layer is a meltblown nonwoven fabric with a filtering effect; the inner layer is a spunbond nonwoven fabric with the function of absorbing moisture(23). Among them, meltblown non-woven fabric serves as the most important component(24). The differences in the process whether electret treatment is performed, weight, and thickness between the two may be the reason why the medical non-woven fabrics are close but fail to meet the standards of surgical masks. The special porous arrangement of medical non-woven fabric enables the steam and other media to penetrate the bag flexibly, which has a significant bacteriostasis effect, and has the characteristics of good breathability, small penetration rate, strong water resistance, and flame retardancy(25). Li Muping et al.(26) found that the double-layer medical non-woven fabric could effectively block bacteria within 3 months. Zou Xiuzhen et al.(27)discovered that the disposable non-woven fabric material had good antibacterial effectiveness, and was consistent with the results of our study. At present, medical non-woven fabrics are usually supplied directly to hospitals by manufacturers or suppliers. Residents can also purchase them through some e-commerce platforms such as Amazon and Taobao, but their quality assurance has yet to be verified.

Although three fabric materials (T-shirt, fleece sweater, and tea towel) tested in our study could not reach the standard of surgical mask in terms of bacterial filtration efficiency, some combinations of the three materials showed a higher level of particle filtration, such as fleece sweater plus hairy tea towel. Studies demonstrated that the filtering performance of fabric materials was similar to surgical masks in some aspects. For example, the permeability of fabric materials such as T-shirts and tea towels under the polydisperse NaCl aerosols was 40% to 90%, while that of a surgical mask was 51% to 89%(15, 16). Davies et al. (12)reported that tea towels demonstrated high filtration efficiency in both Bacillus atrophaeus and MS2 bacteriophage aerosols, and the filtration efficiency of double-layer tea towels was close to that of medical surgical mask. Van der Sande et al.(11) found that the homemade tea towel mask could still play a protective role to a certain degree and would not be affected by supply restrictions, although its protective effect was not as strong as a surgical mask or FFP2 mask. However, the materials of homemade masks are not fully recognized in available literatures, and their performance indexes were not tested and verified systematically. It is not clear whether the protective effects of homemade mask materials meet the relevant national standards. Based on previous studies and expert opinions, our study included as many homemade mask materials with good accessibility as possible, and tested relevant indicators in strict accordance with national standards. We also explicitly reported whether these materials met the standards, which can provide a more scientific reference for the selection of materials for homemade masks.

The study design was to use a layer of material with qualified resistance to surface wetting as the outer layer, a layer of material with qualified particle filtration and bacterial filtration efficiency as the intermediate layer, and a layer of material with better hygroscopicity and skin affinity as the inner layer. Comfort was an important factor that affected the compliance of wearing a mask(28, 29). Results have indicated that materials such as medical gauze and T-shirt, which had fewer internal yam threads, less ventilation resistance, and better skin-friendliness, could be used as inner layer materials. Materials such as non-woven shopping bags and fleece sweater materials, which contained tight surface structure, arranged surface texture in parallel, and better surface wettability, could be used as the outer layer material(30). The filtration performance of the most critical intermediate layer material mainly depended on its fiber characteristics, including diameter, charge and fabric density(12). Only medical non-woven fabrics were close to the standard of medical surgical masks in line with results.

Limitations concerning this study are as follows: the study did not test the flame retardant properties, skin irritation, and delayed-type hypersensitivity of the materials. Samples tested in the study were only the original materials rather than the masks made of these materials. Most of the tested materials were purchased from local supermarkets, thus testing results of these materials could be greatly affected by their types, batches, and manufacturers. The performance of the mask on wearing time, wearing frequency, and environment were not tested because no molded masks were made. All of the data were based on laboratory testing while its actual effectiveness in the protection of the crowd, wearing comfort, adverse reactions still need to be verified by human trials and real-world studies.

## Conclusions

In summary, the study shows that some materials and their combinations for homemade masks could meet several standards of surgical masks. If resources are severely lacking and medical masks cannot be obtained, homemade masks using available materials, based on the results of this study, can minimize the chance of infection to the maximum extent.

## Data Availability

I declare that all data is available.

## Acknowledgments

We thank scientific and technological project supported by West China Hospital of Sichuan University for tacking COVID-19, and also thank technology innovation project supported by Chengdu Science and Technology Bureau for tacking COVID-19.

## Supporting information

**S1 Table. Previous research on homemade mask materials**

**S2 File. Pictures of candidate homemade mask materials**

